# The First Insight into the Hereditary Fusion Gene Landscape of Amyotrophic Lateral Sclerosis

**DOI:** 10.1101/2023.03.14.23287250

**Authors:** Jinfeng Yang, Fenghua Yuan, Anna Palovcak, Ling Fei, Noah Zhuo, NYGC ALS Consortium, Yanbin Zhang, Degen Zhuo

## Abstract

Amyotrophic lateral sclerosis (ALS) is a progressive nervous system disease that causes loss of muscle control. Over 30 mutated genes are associated with ASL. However, 90-95% of ASL cases have been found without a family history. Here, we have analyzed RNA-Seq data of NYGC ALS Consortium and identified fusion transcripts from ASL patients and non-neurologic controls (NNC). In this study, we combined previously-curated 1180 monozygotic (MZ) hereditary fusion genes (HFGs), and 204 HFGs discovered from NNC to analyze ASL fusion transcripts and identified 348 HFGs. Comparative analysis between ASL and GTEx shows that 139 HFGs are associated with ASL and ranged from 10.4% to 98.7% of 77 ASL patients. The most recurrent HFG is *ZNF528-ZNF880*, detected in 98.7% of 77 ASL patients and 4.5% of 133 GTEx brain cortexes. Alignments of HFG transcripts from ASL with fusion transcripts from mesial temporal lobe epilepsy (MTLE) and Alzheimer’s disease (AD) showed that 43.9% and 11.6% of the ASL HFGs were present in MTLE and AD, respectively. The most recurrent and common HFG among ASL, MTLE, and AD was *ADAMTSL3-SH3GL3*, which behaves like ubiquitously-expressed *SH3GL3-ADAMTSL3* epigenetic fusion gene (EFG) and shows that *ADAMTSL3-SH3GL3* is a potential dormant or differentially-expressed HFG (dHFG), suggesting that they have common pathophysiological mechanisms. These HFGs associated with ASL have shown that HFGs are the missing genetic heritability and provide novel therapeutic targets for more efficient therapeutic drugs and methods to treat and cure many neurological diseases.

## Introduction

Amyotrophic lateral sclerosis (ALS) is a progressive, fatal neurodegenerative disease characterized by progressive degenerative changes in upper and lower motor neurons that overlap with frontotemporal lobar degeneration clinically, morphologically and genetically^1^. ASL clinical symptoms include weakness of the bulbar and limb muscles, hyperreflexia, spasticity of the arms and legs, and respiratory failure ^2, 3^. These symptoms start with onset typically in the fifth or sixth decade of life, and death commonly occurs within 2-4 years of diagnosis^2^. ALS is closely related to frontotemporal dementia (FTD), a progressive neurodegenerative disease characterized by degeneration of the frontal and temporal lobes of the brain, resulting in behavior, personality, and language disturbances. Up to 15% of ALS patients are also diagnosed with frontotemporal dementia (FTD-ALS patients)^4^. Many studies have shown clinical, pathological, and genetic commonalities between them. Therefore, ASL and FTD have now considered two manifestations of one disease continuum of the FTD-ALS spectrums^5^.

ALS is associated with over 30 mutated genes, including *SOD1*, *TARDBP*, *FUS*, *OPTN*, VCP, UBQLN2, C9ORF72, and PFN1^6–8^. However, they are typically present in fewer than 5-10% of all ALS patients, and most ASL cases remain unexplained^9 10^. Genome-wide associations (GWAS) have genotyped numerous single nucleotide polymorphisms (SNPs) present in at least 1% of the human population^11^ and identified some common and rare SNPs associated with ASL across the genome ^12–14^. More recently, the discovery of genes associated with ASL has been shifted to next-generation sequencing, such as whole-genome sequencing (WGS) and RNA-Seq. Many WGS studies have confirmed previous findings and identified many common and rare variants associated with ASL ^15, 16^. Brown et al. have performed RNA-Seq analysis on the ASL postmortem cortex samples of NYGC ALS Consortium and found that TDP-43 loss resulted from retrotransposon and ALS-risk SNPs drive mis-splicing and depletion of UNC13A ^17, 18^. Many studies have speculated that structural variants may be a source of missing ASL heritability ^10, 19^. More recently, Raghav et al. analyzed RNA-Seq data from Target ALS and the NYGC ALS Consortium and identified 607 unique fusion genes, some significantly higher in ASL samples than those in control ^20^, suggesting that fusion genes generated by structural variants may be responsible for ASL.

As discussed above, advances in RNA-Seq have made it possible to discover fusion transcripts generated by structural variants. However, misconceptions and poor algorithms for repetitive sequences have made fusion transcripts discovered incomplete, irreproducible, and unreliable ^21^. In order to overcome these problems, we have used the splicingcode theory to develop SCIF (SplicingCodes Identify Fusion Genes) to discover more reproducible and accurate fusion transcripts and make it possible to compare and analyze data among different experiments^22^. Validation of highly-recurrent fusion genes has cast doubts that fusion genes are cancerous and generated from somatic genomic abnormalities. *KANSARL* (*KANSL1-ARL17A*) is validated as the first predisposition (hereditary) fusion genes specific to 29% of populations of European ancestry ^22^. *TPM4-KLF2*, detected in 92.2% of 727 multiple myeloma patients and initially thought to be a multiple myeloma-specific somatic fusion gene, has been detected in all five healthy bone marrows^23^. Based on their genomic structures, genetics, and evolution, we have classified the fusion genes into somatic, epigenetic, and hereditary^24^. Hereditary fusion genes (HFGs) are the fusion genes that offspring inherit from their parents, excluding readthrough transcripts of two identical-stand neighboring genes^25^, named epigenetic fusion genes (EFGs) ^24^. Furthermore, we have used 1180 HFGs identified from monozygotic (MZ) twins to analyze fusion transcripts from multiple myeloma and acute myeloid leukemia (AML) and discovered that HFGs are dominant inherited factors and mutated genes have played minor roles in cancer inheritance ^26, 27^. EFGs have reflected developmentally interactive consequences between genetic and environmental factors^23, 27^.

In this study, we analyzed RNA-Seq data of NYGC ALS Consortium and identified large numbers of fusion transcripts from RNA-Seq data of ASL patients. Surprisingly, we have observed large numbers of highly-recurrent fusion genes from their non-neurological controls, 204 of which are determined to be HFGs. We have used these newly-identified HFGs and the previously-curated 1180 HFGs to analyze the fusion transcripts. From 358 HFGs, we have identified 139 HFGs associated with ASL ranging from 10.4% to 98.7% of ASL patients. We have discovered that many highly recurrent ASL HFGs have also been detected in mesial temporal lobe epilepsy (MTLE) and Alzheimer’s disease (AD), suggesting common pathophysiological mechanisms ^28^. *ADAMTSL3-SH3GL3* HFG behaves like *SH3GL3-ADAMTSL3* EFG, suggesting it has a much higher recurrent frequency than its GTEx brain cortex counterpart. This study has shown that HFGs are the missing heritability of complex diseases and traits and provides potential therapeutic targets for more efficient novel therapeutic drugs and methods for ASL and other neurological diseases.

## Materials and Methods

### Materials

#### Amyotrophic lateral sclerosis RNA-Seq data

Amyotrophic lateral sclerosis (ALS) RNA-Seq data (GEO: GSE124439) was downloaded from NCBI (https://www.ncbi.nlm.nih.gov/bioproject/PRJNA512012). The ASL dataset contained 176 RNA-Seq data and included 145 samples of ASL spectrum from 77 unique ASL patients, 14 samples of other neurological disorders (named as OND to avoid confusion) from seven unique individuals, and 17 non-neurological controls (NNC) from 12 unique individuals. The RNA-Seq was performed on postmortem patients’ frontal cortex, medial motor cortex, and lateral motor cortex.

#### Mesial temporal lobe epilepsy RNA-Seq data

The mesial temporal lobe epilepsy (MTLE) RNA-Seq data (GEO: GSE134697) was downloaded from NCBI (https://www.ncbi.nlm.nih.gov/bioproject/PRJNA556159). The MTLE RNA-Seq dataset contained the hippocampus and neocortex of 17 MTLE patients and two healthy controls. This study used only RNA-Seq data from the neocortex of 17 MTLE patients.

#### Alzheimer’s disease RNA-Seq data

The Alzheimer’s disease (AD) RNA-Seq data (GEO: GSE53697) was downloaded from NCBI (https://www.ncbi.nlm.nih.gov/bioproject/PRJNA232669). The AD data had 17 brain samples, including nine patients and eight controls.

#### GTEx brain cortex RNA-Seq data

the Genotype-Tissue Expression (GTEx) RNA-Seq data (dbGaP: phs000424) was downloaded from NCBI (https://www.ncbi.nlm.nih.gov/bioproject/PRJNA75899). We selected and performed RNA-Seq data from 133 brain cortexes of 133 unique individuals as the healthy controls.

### Methods

#### Identification of fusion transcripts from RNA-Seq data

We used SCIF (SplicingCodes Identify Fusion Genes) to perform RNA-Seq analysis at the default conditions ^22^. Since SCIF used raw RNA-Seq reads without any assembly to discover the fusion junctions and followed by extended sequence alignments, each RNA-Seq read was either a fusion transcript or not a fusion transcript. Human was not involved in determining whether a sample with a fusion junction or a sample with three fusion junctions was hereditary fusion gene-positive. However, one of the worst defects in discovering the hereditary fusion genes from RNA-Seq data was that levels of gene expression, RNA-Seq qualities, and the number of RNA-Seq reads would affect the analysis. Consequently, we must take caution to handle and analyzing data.

#### Statistical analysis

To compare two different populations, we have used the two-tailed Z score analyses to determine whether the two populations differ significantly in their genetic characteristics. We set the null hypothesis to be that there is no difference between the two population proportions. Z scores are calculated based on the following formula:

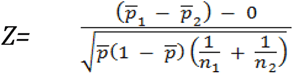

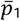 and 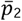 were the population standard errors of p_1_ and p_2_.

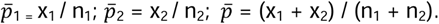

To perform HFG comparisons between ASL and GTEx, if the recurrent frequency was zero in GTEx, we used one to replace zero of the raw count to avoid the infinite.

## Results

### Discovering novel HFGs from RNA-Seq data of the non-neurologic controls

We downloaded the RNA-Seq data of NYGC ALS Consortium (GEO: GSE124439), which contained a total of 176 frontal cortex/motor cortex, including 145 samples of ASL spectrum from 77 unique ASL patients, 14 samples of other neurological disorders (named as OND to avoid confusions) from seven unique individuals, and 17 non-neurological controls (NNC) from 12 unique individuals (Fig.1a). We used SCIF to analyze RNA-Seq data of this dataset and identified 64,000 fusion transcripts from samples of ASL and OND and 8,190 fusion transcripts from 12 non-neurological controls. Manual inspections showed that fusion genes had been discovered at such high recurrent frequencies in non-neurological samples that traditional logic and biological models cannot explain. For example, *ADAMTSL3-SH3GL3*, an inversion of *SH3GL3-ADAMTSL3*, had been observed in all 12 non-neurological controls. If structural variants generating a fusion gene per individual had a rate of 3.6×10^−2^ ^29, 30^, the chance of all 12 NNC individuals having the identical *ADAMTSL3-SH3GL3* fusion gene generated by random somatic genomic alterations was 4.7×10^−18^ and a mathematically impossible event, suggesting it had to be inherited from their parents^24^. Fig.1b shows the procedure to discover hereditary fusion genes (HFGs) from NNC. If we set the cutoff of recurrent frequencies at two or more in 12 NNC individuals, the minimum recurrent frequencies of 16.7% were at least 11.9 fold of 1.4% of the random chance. We initially identified 257 putative HFG transcripts whose recurrent frequencies ranged from 16.7% to 100%. After performing Blast analyses in Aceview and NCBI, we removed 24 EFG transcripts and potential alternative-spliced transcripts and obtained 233 HFG transcripts encoded by 204 HFGs (STable 1). Out of 233 HFG transcripts, we discovered that 33 (16.2%) HFGs coding for 38 transcripts were from tandem gene duplications (STable 2), suggesting that tandem gene duplications were essential for genotype and phenotype diversities^24^.

**Fig. 1.**
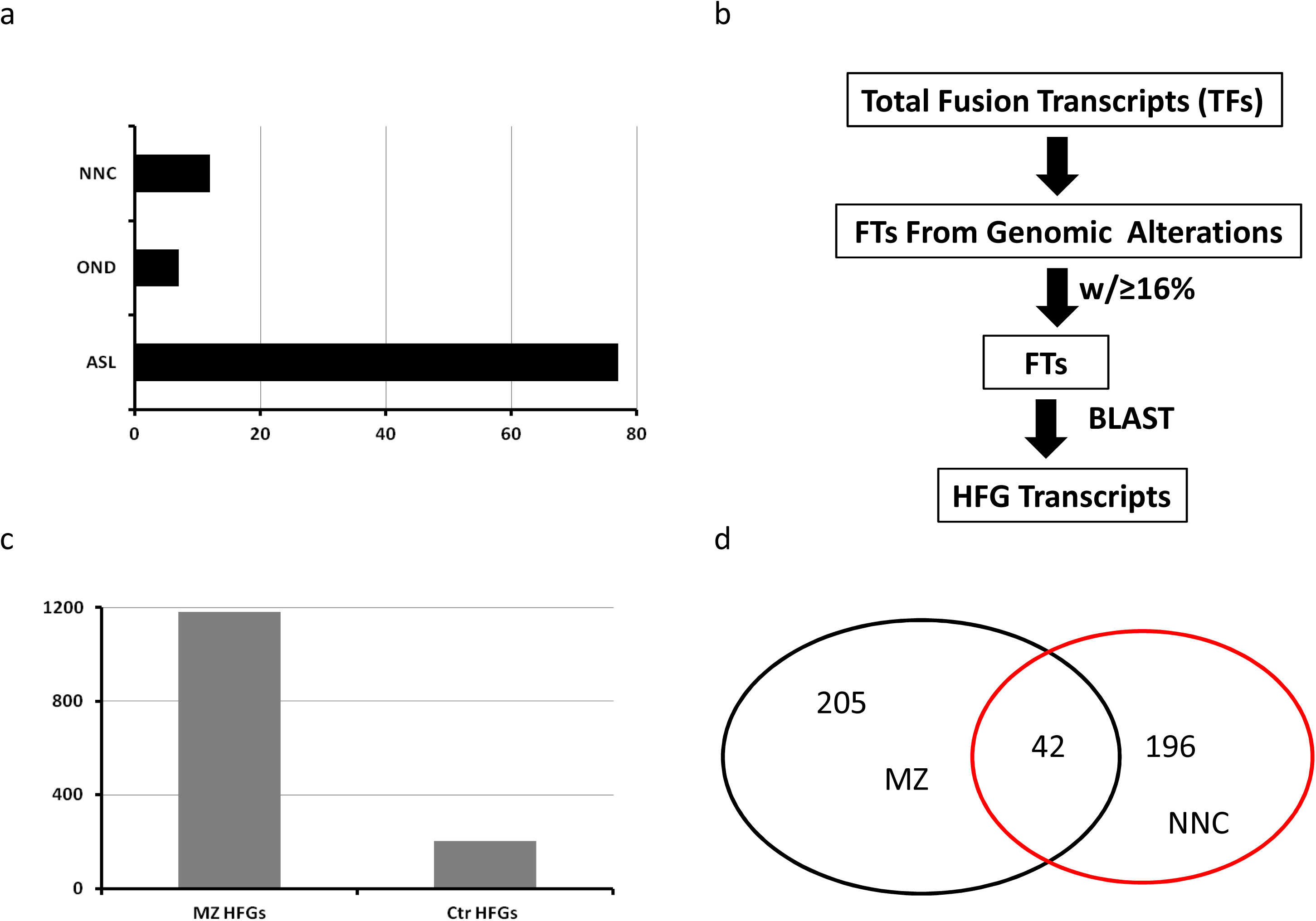
Identification and characterization of hereditary fusion genes (HFGs) from RNA-Seq data of the non-neurological controls (NCC) and amyotrophic lateral sclerosis (ASL). a). characteristics of the amyotrophic lateral sclerosis RNA-Seq dataset (GEO: GSE124439), including 77 amyotrophic lateral sclerosis (ASL) patients, seven patients with other neurological disorders (OND), and 12 non-neurological controls (NNC); b). The procedure of identifying HFGs from 12 NCC individuals; c). The total HFGs used to analyze HFG transcripts of amyotrophic lateral sclerosis, including previously-curated 1180 MZ HFGs and 204 Ctr HFGs; d). The components of HFGs to identify ASL HFGs.

### Identifications of HFGs from RNA-Seq data of amyotrophic lateral sclerosis (ALS) patients

Next, we used these 204 HFGs discovered from NNC plus the previously curated 1180 MZ HFGs^24^ (Fig.1c) to analyze the total fusion transcripts of 154 ASL and 14 OND samples. We identified 358 HFGs encoding 870 HFG transcripts, ranging from 0.6% to 98.1% of 159 ASL and OND samples, the average of which was 2.4 per HFG, suggesting that many HFGs had multiple isoforms. Fig.1d showed that 42 out of 358 HFGs were present in both NNC and MZ HFG datasets and counted for 21.4% of NNC 196 HFGs, supporting our rationales of discovering novel HFGs from NNC. Further analysis showed that only an additional 17 of 196 HFGs were present in GTEx blood samples (data not shown) and suggested that most 196 HFGs were potentially differentially expressed and not expressed in blood and other samples. This differential HFG expression potentially masked the accuracy of the HFG frequencies detected in blood samples. Since these HFG transcripts were from the frontal cortex, medial motor cortex, and lateral motor cortex of ASL and OND samples, we must study whether these HFGs were differentially expressed among the frontal cortex and medial and lateral motor cortexes and affected the results.

To understand whether HFGs were differentially expressed and affected HFG results analyzed afterward, we performed comparative analyses of HFGs among the frontal cortexes, medial motor cortex, and lateral motor cortex. After inspecting the HFG distributions of the frontal cortexes, medial motor cortex, and lateral motor cortexes, we found no HFGs with high recurrent frequencies were differentially expressed among these three brain tissues. STable 3 showed that 179 HFGs were encoding 268 HFG transcripts in all frontal cortex, medial motor cortex, and lateral cortex. Majorities of differences in the HFG transcripts among the three groups were small and statistically insignificant. We identified 22 ASL patients with data from all frontal cortex, medial motor cortex, and lateral motor cortex, similar to 22 three-duplication experiments. We performed a comparative data analysis of these 22 ASL patients. STable 4 showed that we identified 29 HFGs encoding 38 HFG transcripts expressed in all frontal cortex, medial motor cortex, and lateral motor cortex and suggested that these HFGs discovered by SCIF were highly reproducible. For example, *ZNF528-ZNF880* and *SNORD114_-MEG8_* a were observed in all three tissues of 21 of 22 ASL patients and showed 100% reproducibility. Here, *SNORD114_* was an abbreviated *SNORD114-1-SNORD114-2-SNORD114-3*^25^ while *MEG8_* was *MEG8-SNORD112-SNORD113-3*^25^. Careful inspections showed that some HFGs were alternatively spliced. For example, the frequency difference of the main *SNORD114_-MEG8_* isoform a was within statistical errors among the frontal cortex, medial motor cortex, and lateral cortex. However, the recurrent frequencies of the *SNORD114_-MEG8_* isoform c in the medial motor cortex and lateral motor cortexes were 16.4% and 13.7% higher than their counterpart in the frontal cortex. They reflected alternatively splicing in the frontal, medial, and lateral motor cortexes. Since we generally used the recurrent frequency of the main HFG isoform as the HFG recurrent frequency for genotype analysis, we could combine HFG data from the frontal cortex and medial and lateral motor cortexes to perform further analysis.

### Identification and characterization of HFGs associated with ASL

We used the combined HFG dataset described above to analyze the total fusion transcripts from 77 ASL patients. We identified 346 HFGs encoding 819 HFG transcripts whose recurrent frequencies ranged from 1.3% to 98.7%. 345 (42.1%) HFG transcripts were detected in only one ASL patient, suggesting they were either causal HFGs or lowly-expressed HFG transcripts. For a more convenient analysis, we set the cutoff of the recurrent frequency at 10%. STable 5 showed that we observed 147 HFGs coding for 190 HFG transcripts ranging from 10.4% to 98.7%. Table 1 showed that the top sixteen HFGs encoding twenty-one HFG transcripts with recurrent frequencies of ≥50% ranged from 51.3% to 98.7%. Fig.2a showed that the most recurrent *ZNF528-ZNF880*, an inversion of *ZNF880-ZNF528* on the plus strand of chromosome 19q13.41, was detected in 76 (98.7%) of 77 ASL patients and encoded a truncated zinc finger protein 880 without 35 aa N-terminus sequences (S Fig.1). Fig.2b showed that *SNORD114_-MEG8_* was one of the most highly-expressed HFGs and coded for 20 isoforms, three of which were 92.1%, 85.5%, and 53.9% (Table 1). When we performed a comparative analysis between ASL and NCC, many HFG differences were insignificant. For example, *ERMP-KIAA2026*, reported in stomach cancer^31^, was detected in 75% of 75 ASL patients and 12 NNC samples. As described previously, if structural variants generating a fusion gene per individual had a rate of 3.6×10^−2^ ^29, 30^, the chance of having nine NNC individuals who possessed *ERMP-KIAA2026*^31^ by random somatic genomic abnormalities was 1.0×10^−13^ and a mathematically-impossible event. It was most likely that NNC was extraordinarily distorted for some unexplained reasons and unsuitable to use as a standard control.

**Fig. 2.**
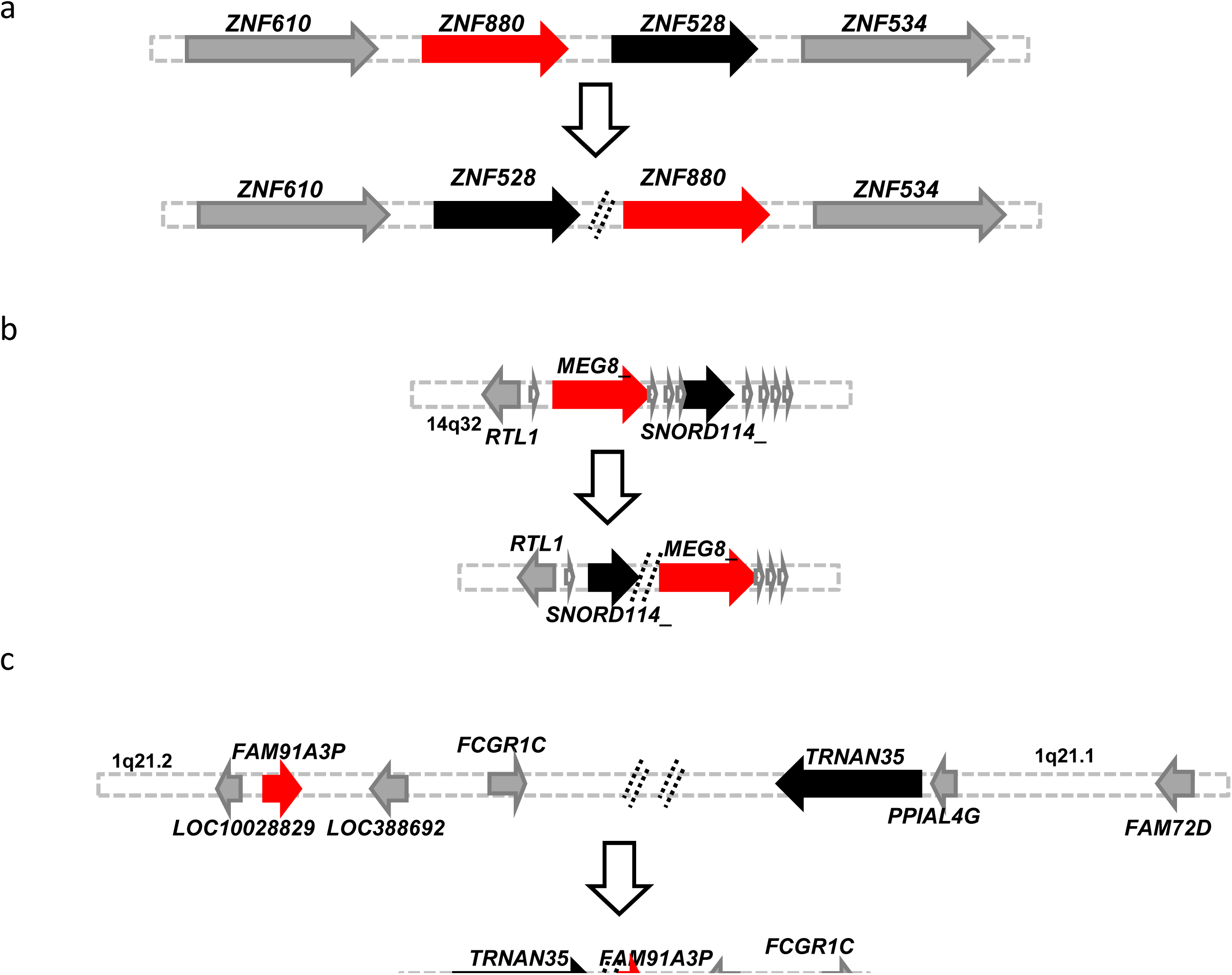
Potential genomic mechanisms to generate *ZNF528-ZNF880*, *TRNAN35-FAM91A3P*, and *SNORD114*_-*MEG8_*. a). A potential local inversion to form *ZNF528-ZNF880*; b) A putative local translocation to produce *TRNAN35-FAM91A3P;* and c). A potential local inversion to generate *SNORD114*_-*MEG8_*, Black and red horizontal arrows indicated 5’ and 3’ genes of HFGs. Gray horizontal arrows were genes surrounding 5’ and 3’ genes. The vertical arrows showed potential genomic rearrangements.

**Table 1.**
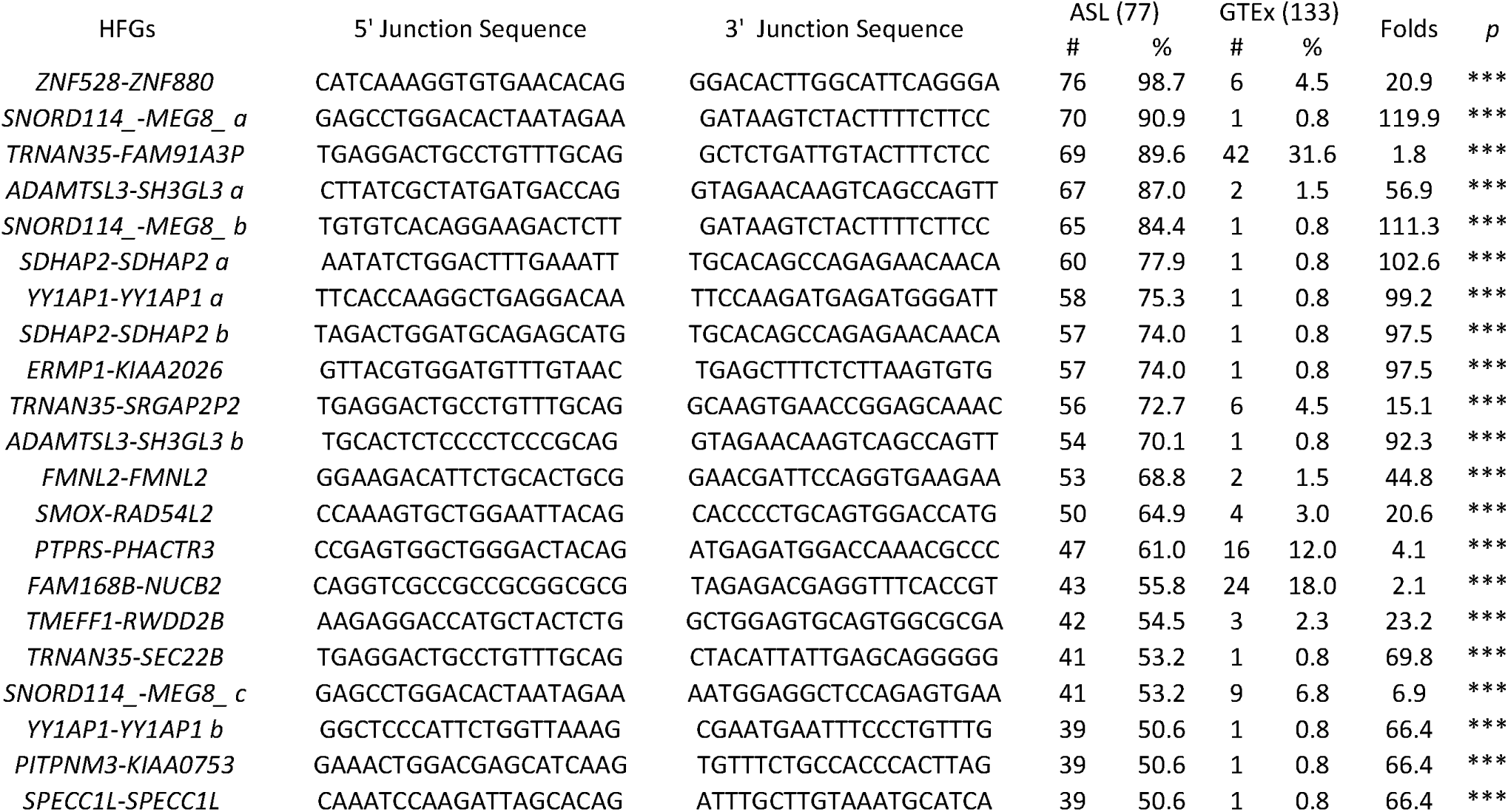
Comparisons of the top sixteen HFGs between ASL and GTEx.

To overcome the NNC distortion, we selected RNA-Seq data from 133 GTEx brain cortexes as healthy controls. We used 346 HFG from 77 ASL patients to analyze 26,000 fusion transcripts discovered from the GTEx brain cortexes and identified 165 HFGs coding for 188 HFG transcripts from GTEx brain cortexes, suggesting that 47.7% of 346 ASL HFGs were present in the GTEx brain cortexes. The most recurrent GTEx HFGs were *KANSARL* (*KANSL1-ARL17A*)^22^ and *TRNAN35-FAM91A3P* (Fig.2c) HFGs were detected in 34.6% and 31.6% of 133 GTEx brain cortexes. In comparison, 56.6% of the ASL HFGs with recurrent frequencies of ≥10% overlapped with HFGs of GTEx brain cortexes, suggesting that the more recurrent ASL HFGs were, the more likely they were present in GTEx brain cortexes.

Next, we performed a comparative analysis of the HFG differences between ASL and GTEx. We discovered that 139 out of 147 ASL HFGs ranged from 10.4% to 98.7% and were 1.2-244 folds higher than their GTEx brain cortex counterparts, which were statistically significant (STable 5). Table 1 showed that sixteen HFGs were encoding twenty-one HFG transcripts, whose recurrent frequencies ranged from 51.3% to 98.7% and were 1.8 to 120 folds of the GTEx brain cortex counterparts, which were statistically significant by the *Z*-test. Table 1 showed that *ZNF528-ZNF880* (Fig.2a) was the most recurrent HFG detected in 98.7% of 77 ASL patients. In comparison, it was detected in 4.5% of 133 GTEx brain cortexes. The former was 21 folds of the latter, suggesting that *ZNF528-ZNF880* HFG was necessary for ASL initiation and prognosis. The most significant difference of HFG recurrent frequencies between ASL and GTEx was *SNORD114_-MEG8_ (*Fig.2b), a local inversion of *SNORD114*-1-*SNORD114*-2-*SNORD114*-3 EFG^25^ and *MEG8-SNORD112-SNORD113-3* EFG^25^, resulted in potentially more diverse expression of snoRNAs. *TRNAN35-FAM91A3P* was the HFG with the least frequency difference between ASL and GTEx. It was an inversion of *TRNAN35*, encoding transfer RNA asparagine 35 and *FAM91A3P*, coding for the family with sequence similarity 91 member A3 pseudogene (Fig.2c), detected in 31.6% of GTEx brain cortexes.

Table 1 showed that *TRNAN35-FAM91A3P*, *TRNAN35-SRGAP2P2*, and *TRNAN35-SEC22B* were three *TRNAN35*-fused HFGs coded for putative 124 a.a, 177 a.a, 171 a.a, and 67 a.a polypeptides, which were potentially regulated by *TRNAN35* via asparaginyl-tRNA synthetase and formed a natural network^24^. Four HFGs, including *SDHAP2-SDHAP2*, *YY1AP1-YY1AP1* (S Fig.2), *FMNL2-FMNL2* (SFig.3), and *SPECC1L-SPECC1L*, were shown to be generated via gene tandem duplications, suggesting that tandem gene duplications were more widespread than what was reported previously^32^. *FMNL2-FMNL2* encoded an FMNL2 protein with two exon insertions. *FMNL2* has been shown to regulate gliovascular interactions and is associated with vascular risk factors and cerebrovascular pathology in Alzheimer’s disease^33^. *YY1AP1-YY1AP1* encoded a truncated YY1-associated protein 1. A genome-wide sequence showed that a frameshift mutation and a large deletion in *YY1AP1* resulted in a girl with a panvascular artery disease^34^. *SPECC1L-SPECC1L* seemed to encode intact SPECC1L protein. It had been shown that mutated *SPECC1L* caused congenital diaphragmatic hernia -- a prominent feature of a *SPECC1L*-related syndrome^35^.

### Identification of common HFGs among ASL, Alzheimer’s disease (AD), and mesial temporal lobe epilepsy (MTLE)

To investigate whether the OND had different HFGs, fusion transcripts identified from 14 OND samples of seven patients were aligned with ASL HFG transcripts. We identified 206 HFGs encoding 318 HFG transcripts whose recurrent frequencies ranged from 14.3% to 100%, suggesting that 59.5% of 346 ASL HFGs were also found in seven OND patients, many of which had much higher recurrent frequencies in OND than in ASL. *SNORD114*_-*MEG8_*, *FMNL2-FMNL2*, and *ADAMTSL3-SH3GL3* were the three top HFGs detected in all seven (100%) OND, while they were observed in 90.1%, 68.8%, and 87% of 77 ASL patients. Many other HFGs associated with ASL were also observed in OND at similar or even higher recurrent frequencies than their ASL counterparts. 59.5% of ASL HFGs observed in OND raised interesting questions about whether other neurological diseases, such as temporal lobe epilepsy (MTLE) and Alzheimer’s disease (AD), had common HFGs.

We used 819 HFG transcripts to analyze fusion transcripts of nine advanced AD patients ^36^ and 17 neocortex samples of seventeen MTLE patients produced for a 2019 failed NIH grant application. We discovered that 152 HFGs coding for 214 HFG transcripts overlapped between ASL and MTLE and counted for 43.9% of 346 HFGs discovered in ASL, suggesting that ASL and MTLE had significantly inherited genetic similarity. In comparison, when 817 HFG fusion transcripts were aligned with fusion transcripts from nine AD patients, we identified only 40 HFGs encoding 74 HFG transcripts, accounting for only 11.6% of 346 HFGs observed in ASL and suggested that AD was much more divergent from ASL than MTLE.

Table 2 showed comparisons of highly-recurrent sixteen HFGs between ASL, MTLE, and AD, while the GTEx brain cortex counterparts were used as controls. All 21 HFG transcripts of sixteen ASL HFGs were present in seventeen MTLE patients. Surprisingly, The *TRNAN35-FAM91A3P*, *TRNAN35-SRGAP2P2*, and *TRNAN35-SEC22B* recurrent frequencies in ASL were 2.3 to 3.8 folds of the MTLE counterparts. The *TRNAN35-FAM91A3P* recurrent frequency of MTLE patients was 23.5% and lower than 31.6% of GTEx brain cortexes (Table 2).

**Table 2.** Distributions of the top sixteen HFGs among OND, MTLE, and AD.

| HFGs | ASL (77) |  | OND (7) |  | MTLE (17) |  | AD (9) |  |
| --- | --- | --- | --- | --- | --- | --- | --- | --- |
|  | # | % | # | % | # | % | # | % |
| *ZNF528-ZNF880* | 76 | 98.7 | 6 | 85.7 | 16 | 94.1 | 8 | 88.9 |
| *SNORD114_-MEG8_ a* | 70 | 90.9 | 5 | 71.4 | 14 | 82.4 | 5 | 55.6 |
| *TRNAN35-FAM91A3P* | 69 | 89.6 | 5 | 71.4 | 4 | 23.5 | 4 | 44.4 |
| *ADAMTSL3-SH3GL3 a* | 67 | 87.0 | 7 | 100.0 | 14 | 82.4 | 9 | 100.0 |
| *SNORD114_-MEG8_ b* | 65 | 84.4 | 7 | 100.0 | 13 | 76.5 | 5 | 55.6 |
| *SDHAP2-SDHAP2 a* | 60 | 77.9 | 4 | 57.1 | 14 | 82.4 | 6 | 66.7 |
| *YY1AP1-YY1AP1 a* | 58 | 75.3 | 6 | 85.7 | 12 | 70.6 | 0 | 0.0 |
| *SDHAP2-SDHAP2 b* | 57 | 74.0 | 4 | 57.1 | 9 | 52.9 | 3 | 33.3 |
| *ERMP1-KIAA2026* | 57 | 74.0 | 5 | 71.4 | 4 | 23.5 | 4 | 44.4 |
| *TRNAN35-SRGAP2P2* | 56 | 72.7 | 4 | 57.1 | 5 | 29.4 | 4 | 44.4 |
| *ADAMTSL3-SH3GL3 b* | 54 | 70.1 | 6 | 85.7 | 9 | 52.9 | 4 | 44.4 |
| *FMNL2-FMNL2* | 53 | 68.8 | 7 | 100.0 | 9 | 52.9 | 0 | 0.0 |
| *SMOX-RAD54L2* | 50 | 64.9 | 3 | 42.9 | 7 | 41.2 | 0 | 0.0 |
| *PTPRS-PHACTR3* | 47 | 61.0 | 5 | 71.4 | 1 | 5.9 | 0 | 0.0 |
| *FAM168B-NUCB2* | 43 | 55.8 | 3 | 42.9 | 4 | 23.5 | 0 | 0.0 |
| *TMEFF1-RWDD2B* | 42 | 54.5 | 6 | 85.7 | 5 | 29.4 | 0 | 0.0 |
| *TRNAN35-SEC22B* | 41 | 53.2 | 5 | 71.4 | 4 | 23.5 | 0 | 0.0 |
| *SNORD114_-MEG8_ c* | 41 | 53.2 | 5 | 71.4 | 5 | 29.4 | 0 | 0.0 |
| *YY1AP1-YY1AP1 b* | 39 | 50.6 | 4 | 57.1 | 4 | 23.5 | 0 | 0.0 |
| *PITPNM3-KIAA0753* | 39 | 50.6 | 3 | 42.9 | 4 | 23.5 | 2 | 22.2 |
| *SPECC1L-SPECC1L* | 39 | 50.6 | 6 | 85.7 | 8 | 47.1 | 0 | 0.0 |

Table 2 also showed that only half of the top sixteen highly recurrent ASL HFGs were detected in AD patients, while another half was absent in AD patients. The most notable HFGs absent in AD patients were *YY1AP1-YY1AP1*, *FMNL2-FMNL2*, and *SPECC1L-SPECC1L* generated by tandem duplications. The most critical HFGs shared between ASL and AD were *ZNF528-ZNF880* and *ADAMTSL3-SH3GL3*, detected in 98.7% and 87% of 77 ASL patients, and 88.9%, and 100% of nine AD patients, respectively. These highly recurrent HFGs were present in ASL, MTLE, and AD and supported that many neurological diseases might have common pathophysiological mechanisms ^28^.

### Identification of *ADAMTSL3-SH3GL3* as a potentially dormant or differentially-expressed HFG

*ADAMTSL3-SH3GL3*, reported recently ^20, 37^, was an inversion of *SH3GL3-ADAMTSL3*^38^ on 15q25, encoding thirteen isoforms and coded for 42 alternatively-spliced isoforms (Fig.3). Its *ADAMTSL3-SH3GL3* main isoform a was expressed only at 19% of the *KANSARL (KANASL1-ARL17A)* main isoform level and encoded a conceptual 202 aa ADAMTSL3-SH3GL3 fusion protein over two-thirds of which was from *SH3GL3*. Since *SH3GL3* was most expressed only in the brain and testis while *ADAMTSL3* was expressed at a minimum level in the brain but at high levels in other tissues, *ADAMTSL3-SH3GL3* may alter *SH3GL3* expression patterns and regulations. Table 3 showed that *ADAMTSL3-SH3GL3* was also detected in ASL, OND, AD, and MTLE and ranged from 82.4% to 100%. Surprisingly, *ADAMTSL3-SH3GL3* was also detected in 100% of 12 NNC samples and 87.5% of eight AD-healthy controls, significantly higher than 1.5% of 133 GTEx brain cortexes (Table 3). To address these enormous gaps among these controls, we used *SH3GL3-ADAMTSL3*, recently reported^38^, as an example to compare both HFG expression patterns and behaviors. Table 3 showed that *SH3GL3-ADAMTSL3*, coding for an intact ADAMTS-like 3 protein, was ubiquitously expressed in all datasets analyzed except for GTEx brain cortexes, in 7.5% of which it was detected. Since the *SH3GL3-ADAMTSL3* locus was present in almost all human individuals, its expression might have been induced due to interactions between environmental and genetic lesions during aging. Hence, we speculated that the *ADAMTSL3-SH3GL3* frequency in human populations was higher than 1.5%. Most *ADAMTSL3-SH3GL3* was dormant or differentially expressed at younger ages and activated by interactions between environmental and genetic lesions during aging. We named this dormant or differentially expressed fusion gene inherited from their parents the dormant or differentially-expressed HFG (dHFG). Hence, *ADAMTSL3-SH3GL3* was the first potential dHFG. Table 3 showed that *SH3GL3-ADAMTSL3* and *ADAMTSL3-SH3GL3* were early-stage aging and neurological disease biomarkers.

**Fig. 3.**
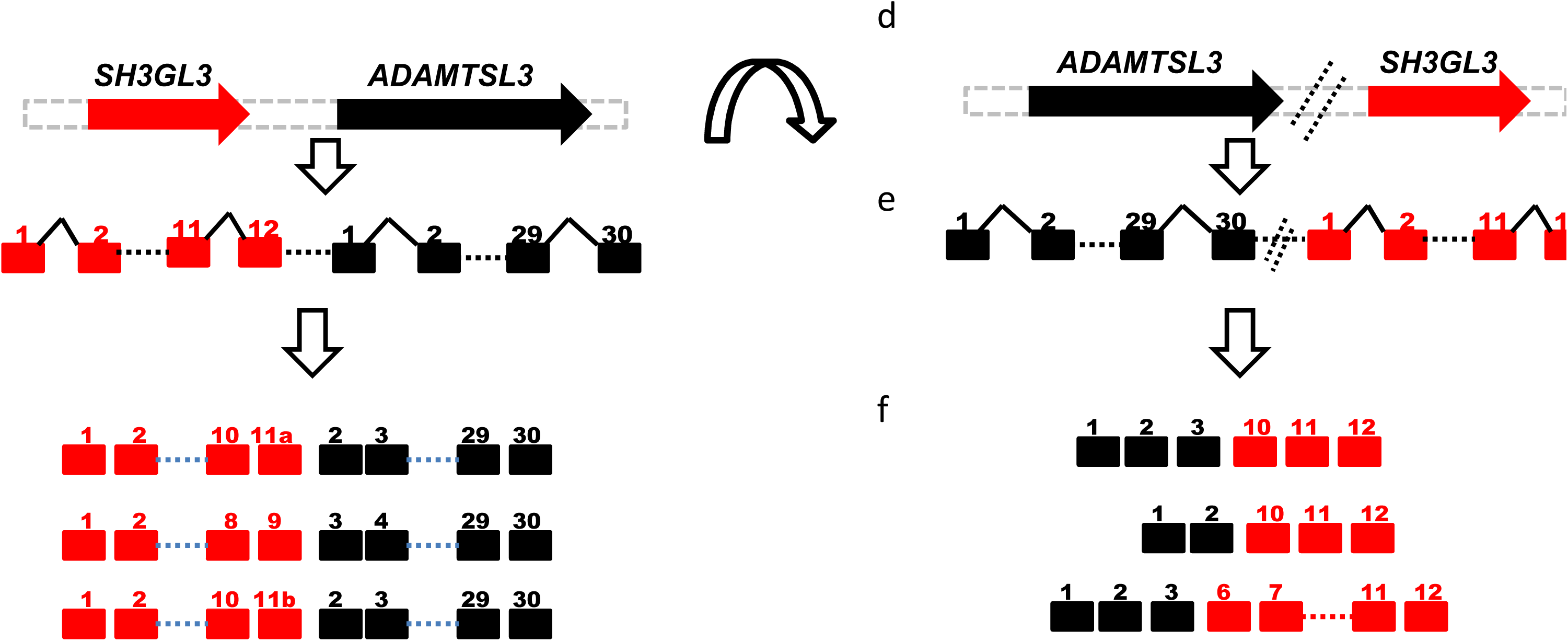
A potential local inversion of *SH3GL3-ADAMTSL3* to produce *ADAMTSL3-SH3GL3*. Two identical-stand neighboring *SH3GL3* and *ADAMTSL3* genes (a) produced an *SH3GL3-ADAMTSL3* readthrough pre-mRNA (b), which was spliced to form three of thirteen *SH3GL3-ADAMTSL3* EFG transcripts(c). *SH3GL3-ADAMTSL3* was locally inverted or locally amplified to *ADAMTSL3-SH3GL3* hereditary fusion gene (HFG) (d), transcribed into *ADAMTSL3-SH3GL3* pre-mRNAs €, which was spliced into three examples of 42 *ADAMTSL3-SH3GL3* HFG transcripts. Black and red horizontal solid arrows showed 5’ and 3’ genes of HFGs. Black and red rectangles were exons. Black triangles were introns. The double dashed lines were putative breakpoints. Horizontal dashed lines indicated omitted exons and introns. An open arch arrow indicated a local translocation or genomic amplification.

**Table 3.**
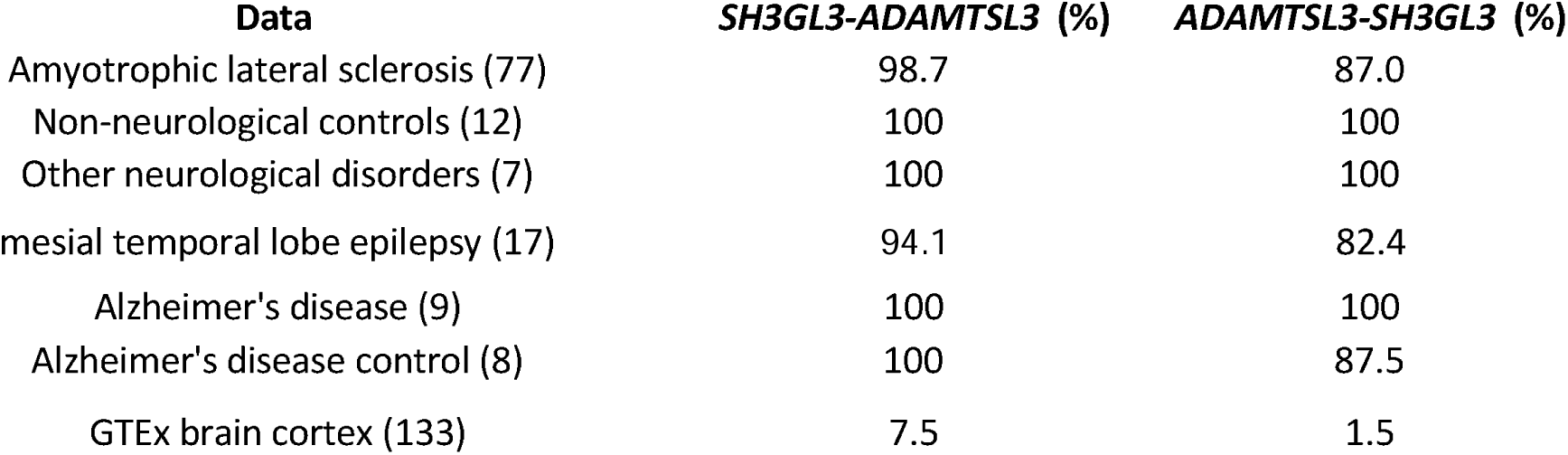
ADAMTSL3-SH3GL3 behaved like SH3GL3-ADAMTSL3 and was a potential dormant HFG.

| **Data** | ***SH3GL3-ADAMTSL3* (%)** | ***ADAMTSL3-SH3GL3* (%)** |
| --- | --- | --- |
| Amyotrophic lateral sclerosis (77) | 98.7 | 87.0 |
| Non-neurological controls (12) | 100 | 100 |
| Other neurological disorders (7) | 100 | 100 |
| mesial temporal lobe epilepsy (17) | 94.1 | 82.4 |
| Alzheimer's disease (9) | 100 | 100 |
| Alzheimer's disease control (8) | 100 | 87.5 |
| GTEx brain cortex (133) | 7.5 | 1.5 |

## Discussion

We analyzed the RNA-Seq data of the NYGC ALS Consortium and identified fusion transcripts from ASL, OND, and NNC. Many NNC fusion transcripts were highly recurrent. For example, *ADAMTSL3-SH3GL3* was detected in 100% of 12 NNC samples and is consistent with our previous finding that *ADAMTSL3-SH3GL3* was detected in 87.5% of eight AD-healthy controls^36^. If structural variants generating a fusion gene per individual had a rate of 3.6×10^−2^ ^29, 30^, generating seven and twelve *ADAMTSL3-SH3GL3* simultaneously in a dataset was 7.8%10^−11^ and 4.7%10^−18^. Such low chances from two different sources^18, 36^ made it mathematically impossible for *ADAMTSL3-SH3GL3* to be generated via random somatic genomic abnormalities. These observations supported that fusion transcripts generated via genomic alterations in healthy controls were potential HFGs. Hence, we developed a novel approach using fusion transcripts of NNC to discover HFGs. We used a mathematical model and blast against AceView and NCBI and identified 204 HFGs encoding 233 HFG transcripts. Bioinformatic analysis showed that 33 (16.2%) HFGs were from tandem gene duplications. As we described previously, SCIF was not intended to discover fusion transcripts generated via tandem duplications, and their discovery was due to gene duplications and pseudogenes^24^. Hence, HFGs produced via genomic tandem duplications were essential biomarkers for diseases and complex traits and might be significantly underestimated.

We combined 204 HFGs discovered from NNC and the previously-curated 1180 MZ HFGs into the HFG dataset to analyze the fusion transcripts of 154 RNA-Seq data from 77 ASL patients. We discovered 346 HFGs encoding 819 HFG transcripts from ASL patients, whose recurrent frequencies ranged from 1 (1.3%) to 76 (98.7) of 77 ASL patients. Out of 819 HFG transcripts, 345 (42.1%) HFG transcripts were detected in only one of 77 ASL patients and were shown to be causal HFGs and lowly-expressed HFG transcripts. When a cutoff of recurrent frequency was set at 10%, we identified 147 HFGs coding for 188 HFG transcripts ranging from 10.4% to 98.7%. As described above, HFGs discovered from 12 NNC samples were extremely recurrent, some were detected in all 12 NNC samples, and the NNC genotypes were distorted. So, we have yet to discover any reasons which can explain these HFG genotype distortions. Hence, we selected 133 GTEx brain cortexes as healthy controls. Table 1 showed that 139 HFGs were 1.2-121 folds higher than their GTEx brain cortex counterparts and were shown by statistical analysis to be associated with ASL. Out of 139 HFGs associated with ASL, Table 2 showed sixteen HFGs with recurrent frequencies of ≥50%, some of which were also highly recurrent in OND, AD, and MTLE. 139 HFGs associated with ASL were four folds of more than 30 mutated genes associated with ASL. The HFG numbers and their recurrent frequencies suggested that HFGs were much more critical “inherited” genetic factors than the mutated genes^6–8^. Hence, we can conclude that highly recurrent HFGs are the missing ASL heritability. More importantly, many neurological diseases such as ASL, MTLE, and AD shared highly recurrent HFGs and supported that these neurological diseases had common pathophysiological mechanisms^28^. They potentially allowed us to develop more effective novel therapeutic drugs and methods to treat and cure neurological diseases.

One of the most highly-recurrent HFGs was *ADAMTSL3-SH3GL3*, an inversion of *SH3GL3-ADAMTSL3* EFG (Fig.3). *ADAMTSL3-SH3GL3* HFG was detected in at least 82.4% of ASL, OND, AD, and MTLE samples. More surprisingly, it was detected in 100% and 87.5% of 12 NNC samples and nine AD controls. Even though OND, AD, and TMLE data sizes are relatively small, the divergent sources of these data compensated for shortcomings. These data suggested that *ADAMTSL3-SH3GL3* would be more recurrent among general populations than 1.5% of GTEx brain cortexes. To address this apparent disparity, we could take a lesson from *SH3GL3-ADAMTSL3* EFG, the ancestors of *ADAMTSL3-SH3GL3.* Table 3 showed that *SH3GL3-ADAMTSL3* was almost ubiquitously expressed in ASL, OND, AD, MTLE, and two controls. In comparison, it was detected in 7.5% of 133 GTEx brain cortexes and suggested that *SH3GL3-ADAMTSL3*^38^ and *ADAMTSL3-SH3GL3*^20, 37^ behaved similarly. Since *SH3GL3-ADAMTSL3* EFG can be treated as a permanent HFG with a recurrent frequency of 100%, we speculated that *ADAMTSL3-SH3GL3* had a much higher recurrent frequency than 1.5% of GTEx brain cortexes and was activated to be expressed by interactions between genetic and environmental lesions during aging. We named this type of HFG the dormant or differentially-expressed HFGs. We needed genotyping general populations to validate if *ADAMTSL3-SH3GL3* was a dHFG or if other factors caused the disparity between neurological disease-related data and GTEx brain cortexes.

To study whether hereditary fusion genes were associated with ASL, AD, MTLE, and other neurological diseases, one of the most critical issues was the accuracy and reproducibility of fusion transcripts discovered from RNA-Seq data. Alternatively, the software systems to discover fusion transcripts from RNA-Seq data were essential to generate accurate and reproducible fusion transcript data. Since more than 20 software systems were available, scientific communities needed more standards to evaluate the qualities of these systems ^39^. In this study, we showed 22 ASL patients had frontal cortex, medial motor cortex, and lateral cortex and were more like three data duplications. It is important to note that we should avoid such a strategy to study epigenetic fusion genes. As shown in STable 2, we identified 29 HFGs encoding 38 HFG transcripts expressed in all three brain tissues. For example, *ZNF528-ZNF880* and *SNORD114_-MEG8_* were observed in the frontal cortex, medial motor cortex, and lateral cortex of 21 of 22 ASL patients and showed 100% reproducibility of fusion transcripts generated by SCIF. Hence, we recommended that scientists and bioinformaticians use these data as reference standards to select a suitable software system for one’s research projects. Since our fusion transcripts could be traced to each RNA-Seq read, one could use fusion junction sequences provided in this study to search the original datasets to validate HFG fusion transcripts, most of which were reproducible except for fusion transcripts generated by DNA duplications.

## Data Availability

GEO: GSE124439
GEO: GSE134697
GEO: GSE53697

## Acknowledges

We appreciate the support from The Target ALS Human Postmortem Tissue Core, the New York Genome Center for Genomics of Neurodegenerative Disease, the Amyotrophic Lateral Sclerosis Association, and the Tow Foundation. We thank the support from the science and technology innovation Program of Hunan Province (Program #: 2021SK4014).

## Supplemental Materials

NYGC ALS Consortium

Hemali Phatnani^5^, Justin Kwan^6^, Dhruv Sareen^7,8^, James R. Broach^9^, Zachary Simmons^10^, Ximena Arcila-Londono^11^, Edward B. Lee^12^, Vivianna M. Van Deerlin^12^, Neil A. Shneider^13^, Ernest Fraenkel^1^, Lyle W. Ostrow^14^, Frank Baas^15,16^, Noah Zaitlen^17^, James D. Berry^18,19^, Andrea Malaspina^19,20,21^, Pietro Fratta^22^, Gregory A. Cox^23^, Leslie M. Thompson^24,25^, Steve Finkbeiner^26^, Efthimios Dardiotis^27^, Timothy M. Miller^28^, Siddharthan Chandran^29^, Suvankar Pal^29^, Eran Hornstein^30^, Daniel J. MacGowan^31^, Terry Heiman-Patterson^32^, Molly G. Hammell^33^, Nikolaos. A. Patsopoulos^34,35^, Oleg Butovsky^36^, Joshua Dubnau^37^, Avindra Nath^38^, Robert Bowser^39,40^, Matthew Harms^41^, Eleonora Aronica^42^, Mary Poss^43^, Jennifer Phillips-Cremins^44^, John Crary^45^, Nazem Atassi^46^, Dale J. Lange^47,48^, Darius J. Adams^49,50^, Leonidas Stefanis^51,52^, Marc Gotkine^53^, Robert H. Baloh^54,55^, Suma Babu^19^, Towfique Raj^56^, Sabrina Paganoni^57^, Ophir Shalem^58,59^, Colin Smith^60,61^, Bin Zhang^62^, Brent Harris^63^, Iris Broce^64^, Vivian Drory^65^, John Ravits^66^, Corey McMillan^67^, Vilas Menon^68^, Lani Wu^69^, Steven Altschuler^69^, Yossef Lerner^70^, Rita Sattler^71^, Kendall Van Keuren-Jensen^72^, Orit Rozenblatt-Rosen^73^, Kerstin Lindblad-Toh^73^, Katharine Nicholson^74^, Peter Gregersen^75^, Jeong-Ho Lee^76^, Sulev Kokos^77^, Stephen Muljo^78^, & Bryan J. Traynor^79^.

^5^Center for Genomics of Neurodegenerative Disease (CGND), New York Genome Center, New York, NY, USA. ^6^Department of Neurology, Lewis Katz School of Medicine, Temple University, Philadelphia, PA, USA. ^7^Cedars-Sinai Department of Biomedical Sciences, Board of Governors Regenerative Medicine Institute and Brain Program, Cedars-Sinai Medical Center, University of California, Los Angeles, CA, USA. ^8^Department of Medicine, University of California, Los Angeles, CA, USA. ^9^Department of Biochemistry and Molecular Biology, Penn State Institute for Personalized Medicine, The Pennsylvania State University, Hershey, PA, USA. ^10^Department of Neurology, The Pennsylvania State University, Hershey, PA, USA. ^11^Department of Neurology, Henry Ford Health System, Detroit, MI, USA. ^12^Department of Pathology and Laboratory Medicine, Perelman School of Medicine, University of Pennsylvania, Philadelphia, PA, USA. ^13^Department of Neurology, Center for Motor Neuron Biology and Disease, Institute for Genomic Medicine, Columbia University, New York, NY, USA. ^1^Department of Biological Engineering, Massachusetts Institute of Technology, Cambridge, MA, USA. ^14^Department of Neurology, Johns Hopkins School of Medicine, Baltimore, MD, USA. ^15^Department of Neurogenetics, Academic Medical Centre, Amsterdam, The Netherlands. ^16^Leiden University Medical Center, Leiden, The Netherlands. ^17^Department of Medicine, Lung Biology Center, University of California, San Francisco, CA, USA. ^18^ALS Multidisciplinary Clinic, Neuromuscular Division, Department of Neurology, Harvard Medical School, Boston, MA, USA. ^19^Neurological Clinical Research Institute, Massachusetts General Hospital, Boston, MA, USA. ^19^Centre for Neuroscience and Trauma, Blizard Institute, Barts, Queen Mary University of London, London, UK. 20 The London School of Medicine and Dentistry, Queen Mary University of London, London, UK. ^21^Department of Neurology, Basildon University Hospital, Basildon, UK. 22 Institute of Neurology, National Hospital for Neurology and Neurosurgery, University College London, London, UK. 23 The Jackson Laboratory, Bar Harbor, ME, USA. ^24^Department of Psychiatry and Human Behavior, Department of Biological Chemistry, School of Medicine, University of California, Irvine, CA, USA. ^25^Department of Neurobiology and Behavior, School of Biological Sciences, University of California, Irvine, CA, USA. ^26^Taube/Koret Center for Neurodegenerative Disease Research, Roddenberry Center for Stem Cell Biology and Medicine, Gladstone Institute, San Francisco, CA, USA. ^27^Department of Neurology and Sensory Organs, University of Thessaly, Thessaly, Greece. ^28^Department of Neurology, Washington University in St Louis, St Louis, MO, USA. ^29^Centre for Clinical Brain Sciences, Anne Rowling Regenerative Neurology Clinic, Euan MacDonald Centre for Motor Neurone Disease Research, University of Edinburgh, Edinburgh, UK. ^30^Department of Molecular Genetics, Weizmann Institute of Science, Rehovot, Israel. ^31^Department of Neurology, Icahn School of Medicine at Mount Sinai, New York, NY, USA. ^32^Center for Neurodegenerative Disorders, Department of Neurology, the Lewis Katz School of Medicine, Temple University, Philadelphia, PA, USA. ^33^Cold Spring Harbor Laboratory, Cold Spring Harbor, NY, USA. ^34^Computer Science and Systems Biology Program, Ann Romney Center for Neurological Diseases, Department of Neurology and Division of Genetics in Department of Medicine, Brigham and Women’s Hospital, Harvard Medical School, Boston, MA, USA. ^35^Program in Medical and Population Genetics, Broad Institute, Cambridge, MA, USA. ^36^Ann Romney Center for Neurologic Diseases, Brigham and Women’s Hospital, Harvard Medical School, Boston, MA, USA. ^37^Department of Anesthesiology, Stony Brook University, Stony Brook, NY, USA. ^38^Section of Infections of the Nervous System, National Institute of Neurological Disorders and Stroke, NIH, Bethesda, MD, USA. ^39^Department of Neurology, Barrow Neurological Institute, St Joseph’s Hospital, Phoenix, AZ, USA. ^40^Medical Center, Department of Neurobiology, Barrow Neurological Institute, St Joseph’s Hospital and Medical Center, Phoenix, AZ, USA. ^41^Department of Neurology, Division of Neuromuscular Medicine, Columbia University, New York, NY, USA. ^42^Department of Neuropathology, Academic Medical Center, University of Amsterdam, Amsterdam, The Netherlands. ^43^Department of Biology and Veterinary and Biomedical Sciences, The Pennsylvania State University, University Park, PA, USA. 44 New York Stem Cell Foundation, Department of Bioengineering, School of Engineering and Applied Sciences, University of Pennsylvania, Philadelphia, PA, USA. ^45^Department of Pathology, Fishberg Department of Neuroscience, Friedman Brain Institute, Ronald M. Loeb Center for Alzheimer’s Disease, Icahn School of Medicine at Mount Sinai, New York, NY, USA. ^46^Department of Neurology, Harvard Medical School, Neurological Clinical Research Institute, Massachusetts General Hospital, Boston, MA, USA. ^47^Department of Neurology, Hospital for Special Surgery, New York, NY, USA. ^48^Weill Cornell Medical Center, New York, NY, USA. 49 Medical Genetics, Atlantic Health System, Morristown Medical Center, Morristown, NJ, USA. 50 Overlook Medical Center, Summit, NJ, USA. ^51^Center of Clinical Research, Experimental Surgery and Translational Research, Biomedical Research Foundation of the Academy of Athens (BRFAA), Athens, Greece. ^52^1^st^ Department of Neurology, Eginition Hospital, Medical School, National and Kapodistrian University of Athens, Athens, Greece. ^53^Neuromuscular/EMG service and ALS/Motor Neuron Disease Clinic, Hebrew University-Hadassah Medical Center, Jerusalem, Israel. ^54^ Board of Governors Regenerative Medicine Institute, Los Angeles, CA, USA. ^55^Department of Neurology, Cedars-Sinai Medical Center, Los Angeles, CA, USA. 56 Departments of Neuroscience, Genetics and Genomic Sciences, Ronald M. Loeb Center for Alzheimer’s disease, Icahn School of Medicine at Mount Sinai, New York, NY, USA. 57 Harvard Medical School, Department of Physical Medicine and Rehabilitation, Spaulding Rehabilitation Hospital, Boston, MA, USA. 58 Center for Cellular and Molecular Therapeutics, Children’s Hospital of Philadelphia, Philadelphia, PA, USA. ^59^Department of Genetics, Perelman School of Medicine, University of Pennsylvania, Philadelphia, PA, USA. ^60^Centre for Clinical Brain Sciences, University of Edinburgh, Edinburgh, UK. ^61^Euan MacDonald Centre for Motor Neurone Disease Research, University of Edinburgh, Edinburgh, UK. ^62^Department of Genetics and Genomic Sciences, Icahn Institute of Data Science and Genomic Technology, Icahn School of Medicine at Mount Sinai, New York, NY, USA. ^63^Department of Neuropathology, Georgetown Brain Bank, Georgetown Lombardi Comprehensive Cancer Center, Georgetown University Medical Center, Washington, DC, USA. ^64^Neuroradiology Section, Department of Radiology and Biomedical Imaging, University of California, San Francisco, San Francisco, CA, USA. ^65^Neuromuscular Diseases Unit, Department of Neurology, Tel Aviv Sourasky Medical Center, Sackler Faculty of Medicine, Tel-Aviv University, Tel-Aviv, Israel. ^66^Department of Neuroscience, University of California San Diego, La Jolla, CA, USA. ^67^Department of Neurology, University of Pennsylvania Perelman School of Medicine, Philadelphia, PA, USA. ^68^Department of Neurology, Columbia University Medical Center, New York, NY, USA. ^69^Department of Pharmaceutical Chemistry, University of California San Francisco, San Francisco, CA, USA. 70 Hadassah Hebrew University, Jerusalem, Israel. ^71^Department of Translational Neuroscience, Barrow Neurological Institute, Phoenix, AZ, USA. 72 The Translational Genomics Research Institute (TGen), Phoenix, AZ, USA. ^73^Broad Institute, Cambridge, MA, USA. ^74^Massachusetts General Hospital, Boston, MA, USA. 75 Institute of Molecular Medicine, Feinstein Institutes for Medical Research, Northwell Health, Manhasset, NY, USA. 76 Korea Advanced Institute of Science and Technology (KAIST), Daejeon, South Korea. ^77^Perron Institute for Neurological and Translational Science, Nedlands, Western Australia, Australia. ^78^Integrative Immunobiology Section, National Institute of Allergy and Infectious Disease, NIH, Bethesda, MD, USA. ^79^Neuromuscular Disease Research Section, National Institute of Aging, NIH, Bethesda, MD, USA

**Supplemental Table 1: Lists of hereditary fusion genes (HFGs) discovered from the non-neurological controls (NNC)**

**Supplemental Table 2: Lists of NNC HFGs formed via tandem gene duplications**

**Supplemental Table 3: Lists of HFGs that were discovered in amyotrophic lateral sclerosis and present in the frontal cortex, medial motor cortex, and lateral motor cortex**

**Supplemental Table 4: Lists and recurrent frequencies of HFGs present in the frontal cortex, medial motor cortex, and lateral motor cortex of 22 ASL patients**

**Supplemental Table 5: Comparative analysis of recurrent frequencies of HFGs between ASL patients and GTEx brain cortexes**

**Supplemental Fig.1 Sequence alignment of conceptual ZNF528-ZNF880 fusion protein and ZNF880 protein. Dashed line indicated the sequence gap.**

**Supplemental Fig.2 Sequence alignment of conceptual FMNL2-FMNL2 fusion protein and formin like 2 protein. Dashed line indicated the sequence gap.**

**Supplemental Fig.3 Sequence alignment of conceptual YY1AP1-YY1AP1 fusion protein and YY1 associated protein 1. Dashed line indicated the sequence gap.**

