## Supplementary material for "The First Insight into the Hereditary Fusion Gene Landscape of Amyotrophic Lateral Sclerosis": . Supplemental Figure 1

ZNF880 and ZNF528-ZNF880 alignment

```
>ZNF880                 MLRRGHLAFRDVAIEFPQEEWKCLDPAQRTLYREVMVENYRNLVFLGICLPDLSVISMLEQ
>ZNF528-ZNF880          -----------------------------------MVENYRNLVFLGICLPDLSVISMLEQ

>ZNF880                 RRDPRNLQSEVKIANNPGGRECIKGVNAESSSKLGSNAGNKSLKNQLGLTFQLHLSELQLF
>ZNF528-ZNF880          RRDPRNLQSEVKIANNPGGRECIKGVNAESSSKLGSNAGNKSLKNQLGLTFQLHLSELQLF

>ZNF880                 QAERNISGCKHVEKPINNSLVSPLQKIYSSVKSHILNKYRNDFDDSPFLPQEQKAQIREKP
>ZNF528-ZNF880          QAERNISGCKHVEKPINNSLVSPLQKIYSSVKSHILNKYRNDFDDSPFLPQEQKAQIREKP

>ZNF880                 CECNEHGKAFRVSSRLANNQVIHTADNPYKCNECDKVFSNSSNLVQHQRIHTGEKPYKCHE
>ZNF528-ZNF880          CECNEHGKAFRVSSRLANNQVIHTADNPYKCNECDKVFSNSSNLVQHQRIHTGEKPYKCHE

>ZNF880                 CGKLFNRISLLARHQRIHTGEKPYKCHECGKVFTQNSHLANHHRIHTGEKPYKCNECGKVF
>ZNF528-ZNF880          CGKLFNRISLLARHQRIHTGEKPYKCHECGKVFTQNSHLANHHRIHTGEKPYKCNECGKVF

>ZNF880                 NRNAHLARHQKIHSGEKPYKCKECGKAFSGGSGLTAHLVIHTGEKLYKCNKCGKVFNRNAH
>ZNF528-ZNF880          NRNAHLARHQKIHSGEKPYKCKECGKAFSGGSGLTAHLVIHTGEKLYKCNKCGKVFNRNAH

>ZNF880                 LTRHQRIHTGEKPYECKECGKVFRHKFCLTNHHRMHTGEQPYKCNECGKAFRDCSGLTAHL
>ZNF528-ZNF880          LTRHQRIHTGEKPYECKECGKVFRHKFCLTNHHRMHTGEQPYKCNECGKAFRDCSGLTAHL

>ZNF880                 LIHTGEKPYKCKECAKVFRHRLSLSNHQRFHTGEKPYRCDECGKDFTRNSNLANHHRIHTG
>ZNF528-ZNF880          LIHTGEKPYKCKECAKVFRHRLSLSNHQRFHTGEKPYRCDECGKDFTRNSNLANHHRIHTG

>ZNF880                 EKPYKCSECHKVFSHNSHLARHRQIHTGEKSYKCNECGKVFSHKLYLKKHERIHTGEKPYR
>ZNF528-ZNF880          EKPYKCSECHKVFSHNSHLARHRQIHTGEKSYKCNECGKVFSHKLYLKKHERIHTGEKPYR

>ZNF880                 CHECGKDFTRNSNLANHHRIHTGEKPYR
>ZNF528-ZNF880          CHECGKDFTRNSNLANHHRIHTGEKPYR
```

Supplemental Fig.1 Sequence alignment of conceptual ZNF528-ZNF880 fusion protein and ZNF880 protein. Dashed line indicated the sequence gap.

FMNL2 and FMNL2-FMNL2  Alignment

```
>FMNL2          MGNAGSMDSQQTDFRAHNVPLKLPMPEPGELEERFAIVLNAMNLPPDKARLLRQYDNEKKW
>FMNL2-FMNL2    MGNAGSMDSQQTDFRAHNVPLKLPMPEPGELEERFAIVLNAMNLPPDKARLLRQYDNEKKW

>FMNL2          ELICDQ------------------------------------------------------
>FMNL2-FMNL2    ELICDQERFQVKNPPHTYIQKLKGYLDPAVTRKKFRRRVQESTQVLRELEISLRTNHIGWV

>FMNL2          ------------------------------------------------------------
>FMNL2-FMNL2    REFLNEENKGLDVLVEYLSFAQYAVTFDFESVESTVESSVDKSKPWSRSIEDLHRGSNLPS

>FMNL2          ----------------ERFQVKNPPHTYIQKLKGYLDPAVTRKKFRRRVQESTQVLRELEI
>FMNL2-FMNL2    PVGNSVSRSGRHSALRERFQVKNPPHTYIQKLKGYLDPAVTRKKFRRRVQESTQVLRELEI

>FMNL2          SLRTNHIGWVREFLNEENKGLDVLVEYLSFAQYAVTFDFESVESTVESSVDKSKPWSRSIE
>FMNL2-FMNL2    SLRTNHIGWVREFLNEENKGLDVLVEYLSFAQYAVTFDFESVESTVESSVDKSKPWSRSIE

>FMNL2          DLHRGSNLPSPVGNSVSRSGRHSALRYNTLPSRRTLKNSRLVSKKDDVHVCIMCLRAIMNY
>FMNL2-FMNL2    DLHRGSNLPSPVGNSVSRSGRHSALRYNTLPSRRTLKNSRLVSKKDDVHVCIMCLRAIMNY

>FMNL2          QYGFNMVMSHPHAVNEIALSLNNKNPRTKALVLELLAAVCLVRGGHEIILSAFDNFKEVCG
>FMNL2-FMNL2    QYGFNMVMSHPHAVNEIALSLNNKNPRTKALVLELLAAVCLVRGGHEIILSAFDNFKEVCG

>FMNL2          EKQRFEKLMEHFRNEDNNIDFMVASMQFINIVVHSVEDMNFRVHLQYEFTKLGLDEYLDKL
>FMNL2-FMNL2    EKQRFEKLMEHFRNEDNNIDFMVASMQFINIVVHSVEDMNFRVHLQYEFTKLGLDEYLDKL

>FMNL2          KHTESDKLQVQIQAYLDNVFDVGALLEDAETKNAALERVEELEENISHLSEKLQDTENEAM
>FMNL2-FMNL2    KHTESDKLQVQIQAYLDNVFDVGALLEDAETKNAALERVEELEENISHLSEKLQDTENEAM

>FMNL2          SKIVELEKQLMQRNKELDVVREIYKDANTQVHTLRKMVKEKEEAIQRQSTLEKKIHELEKQ
>FMNL2-FMNL2    SKIVELEKQLMQRNKELDVVREIYKDANTQVHTLRKMVKEKEEAIQRQSTLEKKIHELEKQ

>FMNL2          GTIKIQKKGDGDIAILPVVASGTLSMGSEVVAGNSVGPTMGAASSGPLPPPPPPLPPSSDT
>FMNL2-FMNL2    GTIKIQKKGDGDIAILPVVASGTLSMGSEVVAGNSVGPTMGAASSGPLPPPPPPLPPSSDT

>FMNL2          PETVQNGPVTPPMPPPPPPPPPPPPPPPPPPPPLPGPAAETVPAPPLAPPLPSAPPLPGT
>FMNL2-FMNL2    PETVQNGPVTPPMPPPPPPPPPPPPPPPPPPPPLPGPAAETVPAPPLAPPLPSAPPLPGT

>FMNL2          SSPTVVFNSGLAAVKIKKPIKTKFRMPVFNWVALKPNQINGTVFNEIDDERILEDLNVDEF
>FMNL2-FMNL2    SSPTVVFNSGLAAVKIKKPIKTKFRMPVFNWVALKPNQINGTVFNEIDDERILEDLNVDEF

>FMNL2          EEIFKTKAQGPAIDLSSSKQKIPQKGSNKVTLLEANRAKNLAITLRKAGKTADEICKAIHV
>FMNL2-FMNL2    EEIFKTKAQGPAIDLSSSKQKIPQKGSNKVTLLEANRAKNLAITLRKAGKTADEICKAIHV

>FMNL2          FDLKTLPVDFVECLMRFLPTENEVKVLRLYERERKPLENLSDEDRFMMQFSKIERLMQKMT
>FMNL2-FMNL2    FDLKTLPVDFVECLMRFLPTENEVKVLRLYERERKPLENLSDEDRFMMQFSKIERLMQKMT

>FMNL2          IMAFIGNFAESIQMLTPQLHAIIAASVSIKSSQKLKKILEIILALGNYMNSSKRGAVYGFK
>FMNL2-FMNL2    IMAFIGNFAESIQMLTPQLHAIIAASVSIKSSQKLKKILEIILALGNYMNSSKRGAVYGFK
```

```
>FMNL2          LQSLDLLLDTKSTDRKQTLLHYISNVVKEKYHQVSLFYNELHYVEKAAAVSLENVLLDVKE
>FMNL2-FMNL2    LQSLDLLLDTKSTDRKQTLLHYISNVVKEKYHQVSLFYNELHYVEKAAAVSLENVLLDVKE

>FMNL2          LQRGMDLTKREYTMHDHNTLLKEFILNNEGKLKKLQDDAKIAQDAFDDVVKYFGENPKTTP
>FMNL2-FMNL2    LQRGMDLTKREYTMHDHNTLLKEFILNNEGKLKKLQDDAKIAQDAFDDVVKYFGENPKTTP

>FMNL2          PSVFFPVFVRFVKAYKQAEEENELRKKQEQALMEKLLEQEALMEQQDPKSPSHKSKRQQQE
>FMNL2-FMNL2    PSVFFPVFVRFVKAYKQAEEENELRKKQEQALMEKLLEQEALMEQQDPKSPSHKSKRQQQE

>FMNL2          LIAELRRRQVKDNRHVYEGKDGAIEDIITDLRNQPYRRADAVRRSVRRRFDDQNLRSVNGA
>FMNL2-FMNL2    LIAELRRRQVKDNRHVYEGKDGAIEDIITDLRNQPYRRADAVRRSVRRRFDDQNLRSVNGA

>FMNL2          EITM
>FMNL2-FMNL2    EITM
```

Supplemental Fig.2 Sequence alignment of conceptual FMNL2-FMNL2 fusion protein and formin like 2 protein. Dashed line indicated the sequence gap.

```
>YY1AP1         MEEEASRSAAATNPGSRLTRWPPPDKREGSAVDPGKRRSLAATPSSSLPCTLIALGLRHEK
>YY1AP1-YY1AP   MEEEASRSAAATNPGSRLTRWPPPDKREGSAVDPGKRRSLAATPSSSLPCTLIALGLRHEK

>YY1AP1         EANELMEDLFETFQDEMGFSNMEDDGPEEEERVAEPQANFNTPQALRFEELLANLLNEQHQ
>YY1AP1-YY1AP   EANELMEDLFETFQDEMGFSNMEDDGPEEEERVAEPQANFNTPQALRFEELLANLLNEQHQ

>YY1AP1         IAKELFEQLKMKKPSAKQQKEVEKVKPQCKEVHQTLILDPAQRKRLQQQMQQHVQLLTQIH
>YY1AP1-YY1AP   IAKELFEQLKMKKPSAKQQKEVEKVKPQCKEVHQTLILDPAQRKRLQQQMQQHVQLLTQIH

>YY1AP1         LLATCNPNLNPEASSTRICLKELGTFAQSSIALHHQYNPKFQTLFQPCNLMGAMQLIEDFS
>YY1AP1-YY1AP   LLATCNPNLNPEASSTRICLKELGTFAQSSIALHHQYNPKFQTLFQPCNLMGAMQLIEDFS

>YY1AP1         THVSIDCSPHKTVKKTANEFPCLPKQVAWILATSKVFMYPELLPVCSLKAKNPQDKILFTK
>YY1AP1-YY1AP   THVSIDCSPHKTVKKTANEFPCLPKQVAWILATSKVFMYPELLPVCSLKAKNPQDKILFTK

>YY1AP1         AEDN-KYLLTCKTARQLTVRIKN----LNMNRAPDNIIKFYKKTKQLPVLGKCCEEIQPHQ
>YY1AP1-YY1AP   AEDNSKMRWDSPTWKMMAQKRRSVWLSLKLTLTP---LKLYGLRNYWPTY-----------

>YY1AP1         WKPPIEREEHRLPFWLKASLPSIQEELRHMADGAREVGNMTGTTEINSDQGLEKDNSELGS
>YY1AP1-YY1AP   -------------------------------------------------------------

>YY1AP1         ETRYPLLLPKGVVLKLKPVADRFPKKAWRQKRSSVLKPLLIQPSPSLQPSFNPGKTPAQST
>YY1AP1-YY1AP   -------------------------------------------------------------

>YY1AP1         HSEAPPSKMVLRIPHPIQPATVLQTVPGVPPLGVSGGESFESPAALPAMPPEARTSFPLSE
>YY1AP1-YY1AP   -------------------------------------------------------------

>YY1AP1         SQTLLSSAPVPKVMMPSPASSMFRKPYVRRRPSKRRGARAFRCIKPAPVIHPASVIFTVPA
>YY1AP1-YY1AP   -------------------------------------------------------------

>YY1AP1         TTVKIVSLGGGCNMIQPVNAAVAQSPQTIPIATLLVNPTSFPCPLNQPLVASSVSPLIVSG
>YY1AP1-YY1AP   -------------------------------------------------------------

>YY1AP1         NSVNLPIPSTPEDKAHMNVDIACAVADGENAFQGLEPKLEPQELSPLSATVFPKVEHSPGP
>YY1AP1-YY1AP   -------------------------------------------------------------
```

```
>YY1AP1         PPVDKQCQEGLSENSAYRWTVVKTEEGRQALEPLPQGIQESLNNSSPGDLEEVVKMEPEDA
>YY1AP1-YY1AP   -------------------------------------------------------------

>YY1AP1         TEEISGFL
>YY1AP1-YY1AP   --------
```

Supplemental Fig.3 Sequence alignment of conceptual YY1AP1-YY1AP1 fusion protein and YY1 associated protein 1. Dashed line indicated the sequence gap.
